# Adverse Outcomes and Mortality in Users of Non-Steroidal Anti-Inflammatory Drugs tested positive for SARS-CoV-2: A Danish Nationwide Cohort Study

**DOI:** 10.1101/2020.06.08.20115683

**Authors:** Lars Christian Lund, Kasper Bruun Kristensen, Mette Reilev, Steffen Christensen, Reimar W. Thomsen, Christian F. Christiansen, Henrik Støvring, Nanna Borup Johansen, Nikolai Brun, Jesper Hallas, Anton Pottegård

## Abstract

**Background:** Concerns over the safety of non-steroidal anti-inflammatory drug (NSAID) use during SARS-CoV-2 infection have been raised.

**Objectives:** To study whether use of NSAIDs is associated with adverse outcomes and mortality during SARS-CoV-2 infection.

**Design:** Population based cohort study

**Setting:** Danish administrative and health registries.

Participants Individuals tested positive for SARS-CoV-2 during Feb 27, 2020 to Apr 29, 2020. Treated individuals (defined as a filled NSAID prescription up to 30 days before the SARS-CoV-2 test) were matched to up to 4 non-treated individuals on propensity scores based on age, sex, relevant comorbidities and prescription fills.

**Outcome measures:** The main outcome was 30-day mortality and treated individuals were compared to untreated individuals using risk ratios (RR) and risk differences (RD). Secondary outcomes included hospitalisation, intensive care unit (ICU) admission, mechanical ventilation and acute renal replacement therapy.

**Results:** A total of 9236 SARS-CoV-2 PCR positive individuals were eligible for inclusion. Of these, 248 (2.7%) had filled a prescription for NSAIDs and 535 (5.8%) died within 30 days. In the matched analyses, treatment with NSAIDs was not associated with 30-day mortality (RR 1.02, 95% CI 0.57 to 1.82; RD 0.1%, −3.5% to 3.7%), increased risk of hospitalisation (RR 1.16, 0.87 to 1.53; RD 3.3%, −3.4% to 10%), ICU-admission (RR 1.04, 0.54 to 2.02; RD 0.2%, −3.0% to 3.4%), mechanical ventilation (RR 1.14, 0.56 to 2.30; RD 0.5%, - 2.5% to 3.6%), or renal replacement therapy (RR 0.86, 0.24 to 3.09; RD −0.2%, −2.0% to 1.6%).

**Conclusion:** Use of NSAIDs was not associated with 30-day mortality, hospitalisation, ICU-admission, mechanical ventilation or renal replacement therapy in Danish individuals tested positive for SARS-CoV-2.

**Registration:** The European Union electronic Register of Post-Authorisation Studies, EUPAS-34734 (http://www.encepp.eu/encepp/viewResource.htm?id=34735)

## Introduction

As the severe acute respiratory syndrome coronavirus 2 (SARS-CoV-2) began to transmit and spread in Europe and the United States, a letter suggesting that ibuprofen could influence the prognosis of coronavirus disease 2019 (COVID-19) through upregulation of angiotensin converting enzyme 2 receptors, was circulated widely (1). Together with case reports of otherwise healthy young patients with severe COVID-19 who had used NSAIDs in the early stage of disease (2), concerns regarding the safety of NSAID use during the COVID-19 pandemic were widely circulated, including warnings against NSAID use in COVID-19 from the French health minister (2) and the World Health Organization. However, the European Medicines Agency stated that evidence to support warnings against use of NSAIDs in COVID-19 was lacking and stressed the need for further evidence on any effects of NSAIDs on disease prognosis in COVID-19 (3).

The available evidence stems mainly from studies on community acquired bacterial pneumonia and show that use of NSAIDs is associated with bacterial complications, specifically empyema and lung abscesses (4–7). For viral illness, use of NSAIDs was not associated with mortality in ICU patients with influenza H1N1 during the 2009 pandemic (8), and a recent study found that use of NSAIDs was not associated with mortality in patients hospitalized for influenza (9). As use of ibuprofen and other NSAIDs is widespread, data on its safety is urgently needed to guide clinicians and patients on how to use NSAIDs during the COVID-19 pandemic. We therefore examined whether use of NSAIDs prior to infection with SARS-CoV-2 was associated with increased risk of hospitalisation, intensive care admission, and mortality compared to non-use of NSAIDs.

## Methods

We conducted a Danish nationwide registry-based cohort study, investigating the association between NSAID use and 30-day mortality and other adverse outcomes, specified as hospitalisation, intensive care unit admission, mechanical ventilation and acute renal replacement therapy in all patients tested positive for SARS-CoV-2. All analyses followed the publicly registered protocol (10), except for a change in the matching algorithm and a *post hoc* analysis of test negative individuals (both detailed below).

### Data sources

Data on all Danish residents with a positive test for SARS-CoV-2 were obtained from Danish health and administrative registries as described elsewhere (11). In brief, identification of the study population was based on prospectively collected data on all Danish residents receiving a polymerase chain reaction (PCR)-test for SARS-CoV-2 from the Danish Microbiology Database (12). Data was linked to the Danish Civil Registration system (13), the Danish National Prescription Registry (14), the Danish National Patient Registry (15), and the Danish Register of Causes of Death (16) by means of the unique personal identifier assigned to all Danish residents at birth or immigration. Data on intensive care unit treatment, mechanical ventilation, and renal replacement therapy from the Danish National Patient Registry was supplemented with daily reports on admitted patients from the five Danish Regions (15,17). For more details regarding individual registries, see supplementary (**Appendix A**).

### Study population

All Danish residents that had a positive polymerase chain reaction (PCR) test for SARS-CoV-2 during the period 27-02-2020 to 29-04-2020 were included in the study. To ensure complete information on exposure and covariates prior to cohort entry, individuals with less than one year of residence in Denmark prior to the positive test for SARS-CoV-2 were excluded. For all secondary outcomes, individuals with an outcome during 30 days to 1 day prior to cohort entry were excluded, partly to ensure that outcomes were incident and plausibly occurring due to COVID-19 and partly because in-hospital drug use was not available from the Danish Prescription Registry.

We conducted a *post-hoc* supplementary analysis where we examined the same association in a cohort of all Danish patients tested negative for SARS-CoV-2 in the study period.

### Exposure

The exposure of interest was current use of any NSAID prior to a positive SARS-CoV-2 test. Current use was defined as having filled a prescription for any NSAID in the 30 days prior to the positive test. Filled NSAID prescriptions were identified from the Danish Prescription Registry with information on all dispensed prescriptions at community pharmacies in Denmark since 1995 (18). Users of NSAID were compared to individuals without NSAID use in the corresponding time window. In Denmark, NSAIDs are sold by prescription except for low-dose (200 mg) ibuprofen sold over-the-counter in pack sizes of no more than 20 tablets. In 2018, over-the-counter purchases of ibuprofen constituted 15% of total ibuprofen sales and a smaller proportion of total NSAID sales (19). Hence, the potential to identify NSAID use from the Danish Prescription Registry is high compared to many other countries where over-the-counter use of NSAIDs is common (18,20).

### Outcomes

The main outcome of interest was death within 30 days of a positive test for SARS-CoV-2. The secondary outcomes included hospitalisation, intensive care unit admission, mechanical ventilation and acute renal replacement therapy within 14 days of a positive test for SARS-CoV-2.

### Follow-up

Eligible individuals were followed until the end of follow up (30 days for the main outcome, 14 days for secondary outcomes) or the event of interest.

### Propensity Score Matching

We used propensity score (PS) matching to increase comparability between NSAID users and non-users. The PS is the estimated probability of receiving the treatment of interest given a set of characteristics (21). Propensity scores were estimated on the day of the positive SARS-CoV-2 test using logistic regression. Independent variables in the PS model were age, sex, relevant comorbidities, use of selected prescription drugs, and phase of the outbreak. For a detailed list of independent variables, see supplementary (**Appendix A**). A separate PS was estimated for each exposure definition in the main and supplementary analyses. To evaluate the appropriateness of the model, PS distributions were plotted separately for each cohort and overlap assessed visually. To reduce unmeasured confounding, individuals in the tails of the PS distribution were trimmed asymmetrically (22). Up to four non-users were matched to each NSAID user using a nearest neighbour algorithm. Non-users could be matched to multiple NSAID users, and the maximum allowed difference in the PS between matches was 0.05 (23). The Danish SARS-CoV-2 test strategy was subject to marked changes from a limited capacity setting in the beginning of the study period to a setting where widespread testing was available at the end of the study period. To account for this (24), we included calendar week of the test-date as a forced matching variable. This decision was made *post-hoc*, i.e. not included in the protocol. Covariate balance before and after matching was assessed using standardized mean differences (25).

### Statistical analyses

Descriptive statistics were used to describe NSAID users and non-users at baseline. Continuous variables were reported as medians and interquartile range. Dichotomous variables were reported as frequencies and percentages.

Risks and risk differences were estimated using generalized linear models with a binomial distribution and an identity link. Risk ratios were estimated similarly but using a log link. Matched analyses were implemented using frequency weighting, i.e. NSAID users were assigned a weight of one, and non-users’ weights were assigned according to each individual user’s number of matches. Robust 95% confidence intervals were calculated using the sandwich estimator of variance where the assumption regarding independence of observations was relaxed in the matched analyses. Data management and statistical analyses were performed using Stata 16 MP. The codes used to define exposures, covariates, and outcomes are available in the supplementary material (**Appendix A**)

### Subgroup analyses

To explore treatment effect heterogeneity, we repeated the main analyses stratifying by age (< 65 years, 65+ years), sex and history of cardiovascular disease. To examine whether widespread testing of health-care workers influenced the findings, we repeated the main analyses restricting the study population to non-health care workers. We used the same propensity score as estimated in the main analyses for the subgroup analyses (26).

### Supplementary analyses

1. We relaxed the exposure definition by using an extended NSAID exposure assessment window prior to the positive test of 60 days and repeated the main analyses with this exposure definition.
2. To explore whether reverse causation may have influenced the findings, effect estimates were obtained using an exposure assessment window of days 60 to days 14 before cohort entry (i.e., disregarded NSAID prescriptions filled during the 14 days immediately prior to cohort entry).
3. To evaluate the robustness of the findings with regards to the chosen outcome assessment windows, we obtained 60-day risk estimates for mortality and 30-day risk estimates for secondary outcomes.
4. To examine the potential for residual confounding, we conducted a negative control analysis by repeating the main analyses within the test negative population, i.e. individuals that were tested negative for SARS-CoV-2 (and not later tested positive). If an individual was tested more than once, the first test date was used as cohort entry date. This *post hoc* analysis was not specified in the protocol.

## Results

We identified 9370 individuals who were tested positive for SARS-CoV-2 during the study period. Of these, 134 were excluded due to migrations within 1 year prior to cohort entry, resulting in an eligible population of 9236 individuals followed for a total of 705 person-years. The median age in the study cohort was 50 years and 58% were female. Overall, 5.8% died within 30 days, 16% were hospitalized within 14 days, 3.1% were admitted to the ICU, 2.5% received mechanical ventilation and 0.7% received acute renal replacement therapy (**Table 1**). 248 (2.7%) patients had filled a prescription for NSAIDs within 30 days before the test date. Compared to non-users, NSAID users were older (median age 55 vs 49 years) and more likely to have a history of hospital-diagnosed overweight or obesity (13% vs 9%) and frequent medical indications for NSAID use such as osteoarthrosis (19% vs 12%), rheumatoid arthritis (7% vs 3%), and to be prescribed opioids the year before sampling date (24% vs 11%). After matching, covariates were well-balanced with standardized mean differences (SMD) ≤ 0.1 (**Table 1**). Use of opioids was strongly associated with use of NSAIDs and 30-day mortality, while cardiovascular disease and dementia was negatively associated with use of NSAIDs and positively associated with death (**Supplementary table S1**).

**Figure 1:**
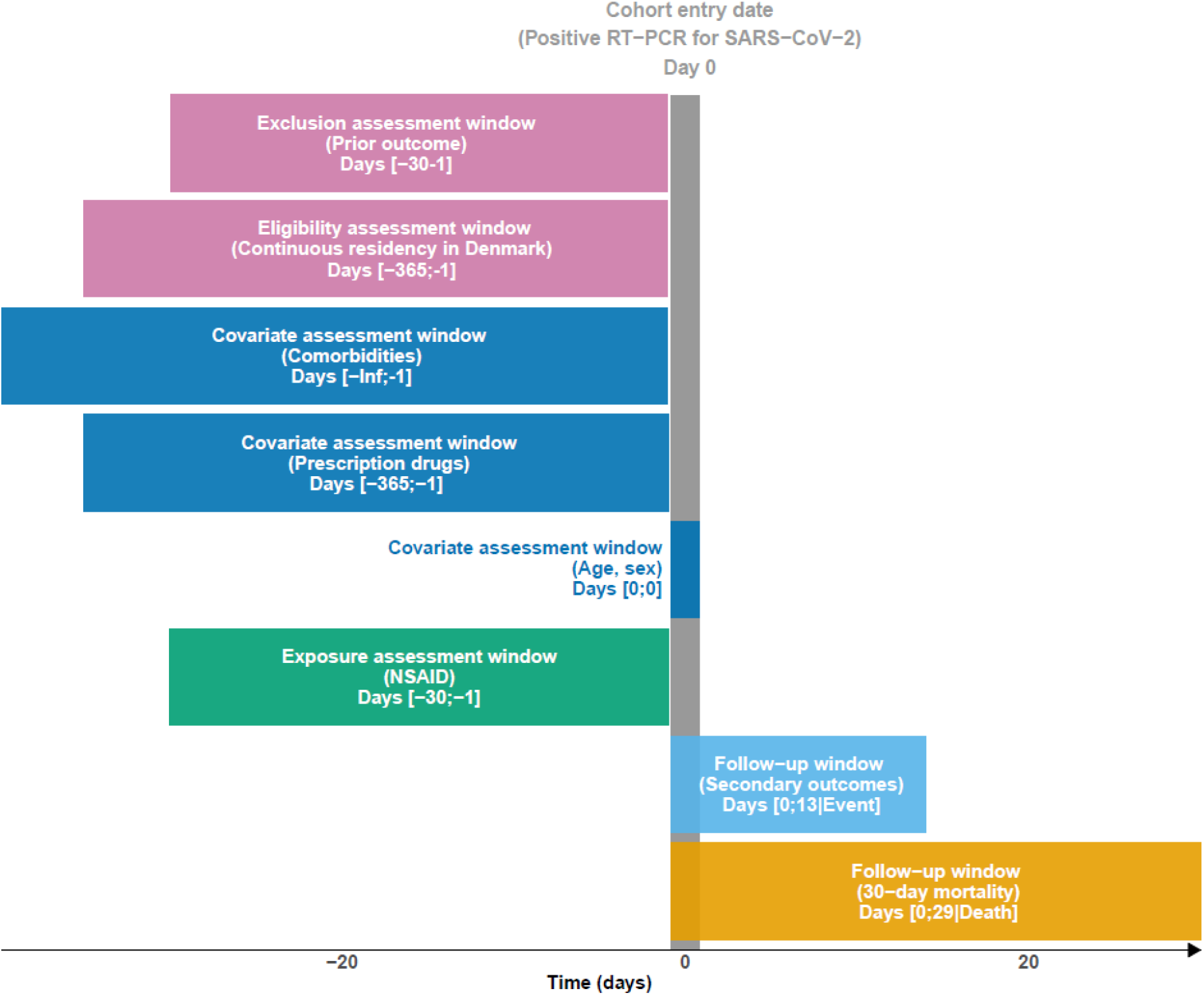
Study design diagram

**Table 1:** Baseline characteristics in the unmatched and propensity score matched cohorts

|  | Unmatched |  |  | Matched |  |  |
| --- | --- | --- | --- | --- | --- | --- |
|  | Treated<br>(n=248) | Untreated<br>(n=8,988) | SMD | Treated<br>(n=224) | Untreated<br>(n=896) | SMD |
| Age, median (IQR) | 55 (43-64) | 49 (35-63) | 0.24 | 54 (43-64) | 54 (41-66) | 0.00 |
| Sex (male) | 99 (39.9) | 3,793 (42.2) | 0.05 | 90 (40.2) | 375 (41.9) | 0.03 |
| Prescriptions* |  |  |  |  |  |  |
| Antihypertensives | 72 (29.0) | 2,221 (24.7) | 0.10 | 62 (27.7) | 233 (26.0) | 0.04 |
| Antidiabetic drugs | 26 (10.5) | 680 (7.6) | 0.10 | 21 (9.4) | 78 (8.7) | 0.02 |
| Low-dose aspirin | 16 (6.5) | 532 (5.9) | 0.02 | 15 (6.7) | 47 (5.2) | 0.06 |
| Immunosuppressants | (n<5) | 63 (0.7) | 0.05 | (n<5) | 6 (0.7) | 0.07 |
| Opioids | 59 (23.8) | 950 (10.6) | 0.36 | 46 (20.5) | 172 (19.2) | 0.03 |
| Z-drugs | 8 (3.2) | 279 (3.1) | 0.01 | 7 (3.1) | 28 (3.1) | 0.00 |
| Benzodiazepines | 10 (4.0) | 378 (4.2) | 0.01 | 10 (4.5) | 38 (4.2) | 0.01 |
| 1st gen. antipsychotics | (n<5) | 58 (0.6) | 0.03 | (n<5) | (n<5) | 0.02 |
| 2nd gen. antipsychotics | (n<5) | 224 (2.5) | 0.10 | (n<5) | 11 (1.2) | 0.03 |
| Systemic glucocorticoids | 19 (7.7) | 431 (4.8) | 0.12 | 15 (6.7) | 65 (7.3) | 0.02 |
| Inhaled corticosteroids | 27 (10.9) | 625 (7.0) | 0.14 | 21 (9.4) | 92 (10.3) | 0.03 |
| Prior diagnoses** |  |  |  |  |  |  |
| Asthma | 16 (6.5) | 613 (6.8) | 0.01 | 13 (5.8) | 47 (5.2) | 0.02 |
| COPD | 11 (4.4) | 368 (4.1) | 0.02 | 9 (4.0) | 35 (3.9) | 0.01 |
| Cardiovascular disease | 28 (11.3) | 1,238 (13.8) | 0.08 | 23 (10.3) | 91 (10.2) | 0.00 |
| Ischaemic stroke | 9 (3.6) | 376 (4.2) | 0.03 | 8 (3.6) | 30 (3.3) | 0.01 |
| Chronic kidney failure | (n<5) | 126 (1.4) | 0.11 | (n<5) | (n<5) | 0.06 |
| Liver disease | (n<5) | 125 (1.4) | 0.02 | (n<5) | 10 (1.1) | 0.06 |
| Alcohol related disorders | 5 (2.0) | 239 (2.7) | 0.04 | (n<5) | 12 (1.3) | 0.04 |
| Dementia | (n<5) | 154 (1.7) | 0.08 | (n<5) | 10 (1.1) | 0.02 |
| Cancer | 21 (8.5) | 646 (7.2) | 0.05 | 16 (7.1) | 64 (7.1) | 0.00 |
| Overweight or obesity | 33 (13.3) | 765 (8.5) | 0.15 | 29 (12.9) | 111 (12.4) | 0.02 |
| Hemiplegia and paraplegia | (n<5) | 35 (0.4) | 0.00 | (n<5) | (n<5) | 0.02 |
| Osteoarthritis | 47 (19.0) | 1,054 (11.7) | 0.20 | 37 (16.5) | 143 (16.0) | 0.02 |
| Rheumatoid arthritis | 17 (6.9) | 308 (3.4) | 0.16 | 13 (5.8) | 51 (5.7) | 0.00 |
| Dysmenorrhoea | 7 (2.8) | 62 (0.7) | 0.16 | (n<5) | 8 (0.9) | 0.00 |

\*Defined as one or more prescription fills during 365 days to 1 day prior to cohort entry
\*\*Defined as one or more discharge diagnoses assigned up to 1 day prior to cohort entry

### Main outcomes

NSAID use was not associated with 30-day mortality in the crude (unmatched) analyses or adjusted (matched) analyses (**Table 2**). In the adjusted analyses, the 30-day mortality was 6.3% in NSAID users and 6.1% in non-users, corresponding to a risk difference of 0.1% (95% CI: −3.5 to 3.7) and a risk ratio of 1.02 (95% CI: 0.57 to 1.82).

**Table 2:**
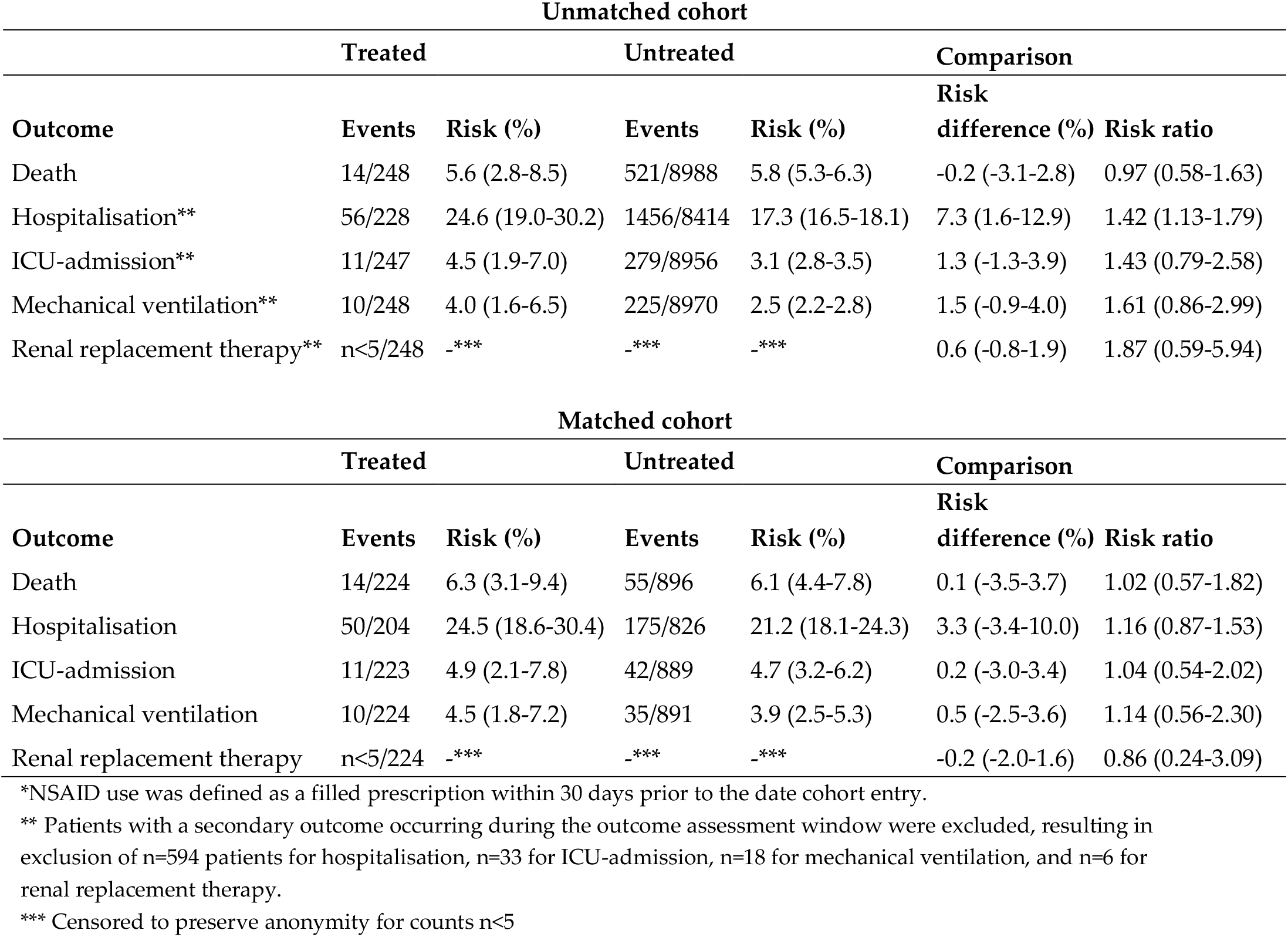
Association between current NSAID use and 30-day mortality, intensive care unit admission, mechanical ventilation, dialysis and hospitalisation in unmatched and propensity score matched cohorts

### Secondary outcomes

In the crude analyses, use of NSAIDs was associated with an increased risk of hospitalisation (RR 1.42, 95% CI: 1.13 to 1.79) and an increased risk of ICU admission (RR 1.43, 95% CI: 0.79 to 2.58), mechanical ventilation (RR 1.61, 95% CI: 0.86 to 2.99), and renal replacement therapy (RR 1.87, 95% CI: 0.59 to 5.94), although these associations were not statistically significant (**Table 2**). After adjustment, NSAID use was not associated with hospitalisation (RR 1.16, 95% CI 0.87 to 1.53), ICU-admission (RR 1.04, 95% CI 0.54 to 2.02), mechanical ventilation (RR 1.14, 95% CI 0.56 to 2.30) or renal replacement therapy (RR 0.86, 95% CI 0.24 to 3.09).

### Subgroup analyses

The subgroup analyses were limited by low power and wide confidence intervals (**Table 3**). In individuals below 65 years of age, the adjusted risk ratio for death associated with NSAID use was 3.76 (95% CI 0.53 to 26.6). After adjustment, use of NSAIDs was associated with hospitalisation in females (RR 1.55, 95% CI: 1.03-2.34) but not in males (RR 0.92, 95% CI 0.62 to 1.36). Otherwise, the risk ratios were not modified by patient characteristics.

**Table 3:**
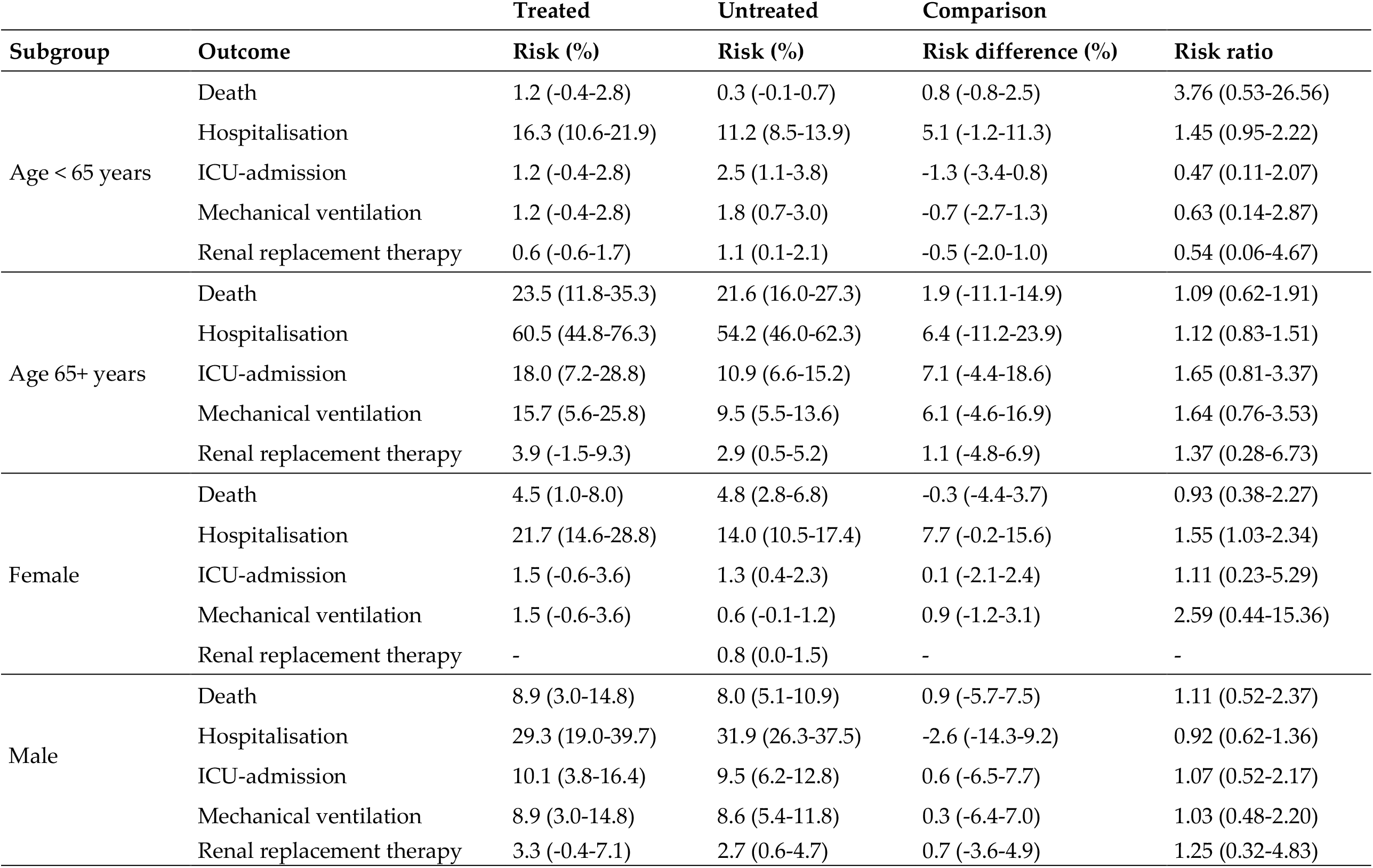
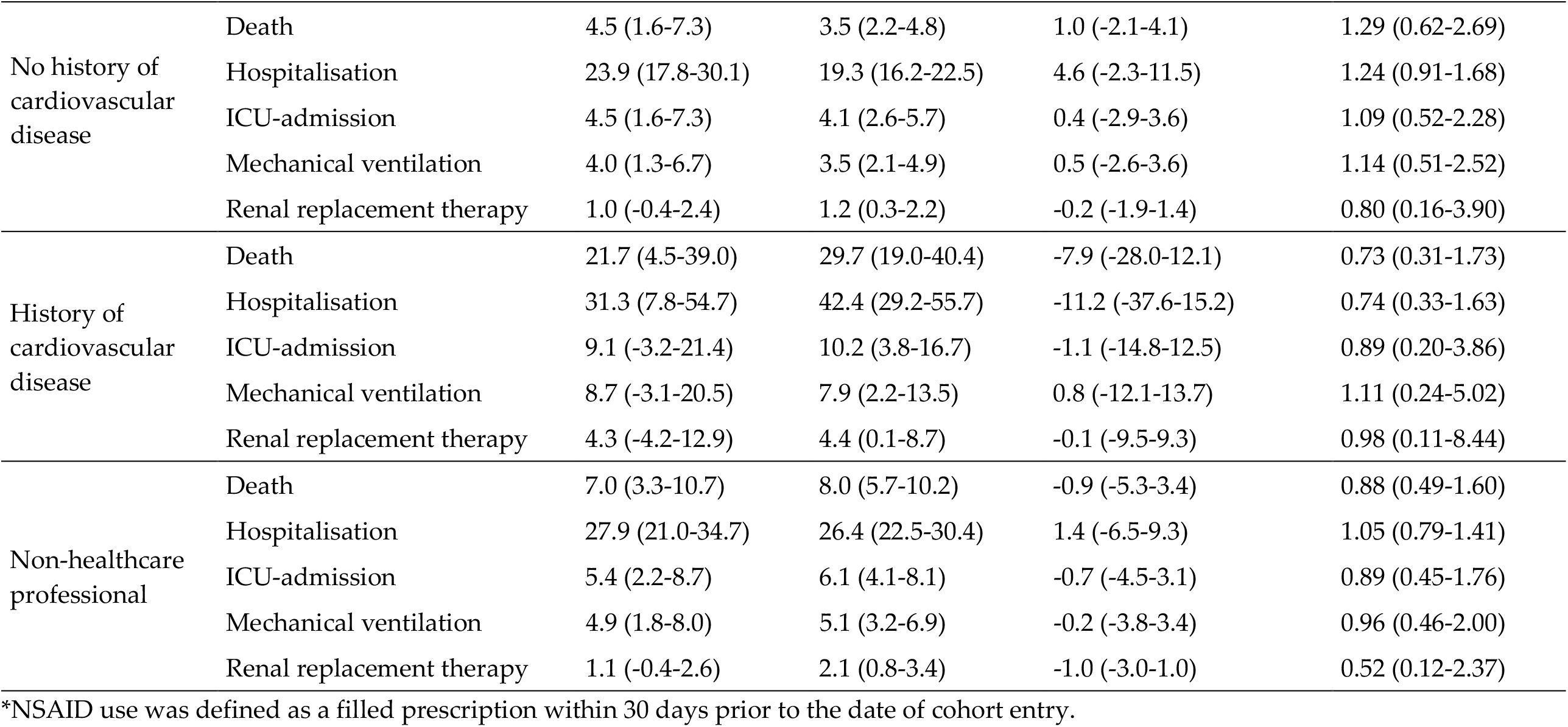
Association between current NSAID use and 30-day mortality, intensive care unit admission, mechanical ventilation, dialysis and hospitalisation in propensity score matched cohorts according to subgroups of interest.

| Subgroup | Outcome | Treated | Untreated | Comparison |  |
| --- | --- | --- | --- | --- | --- |
|  |  | Risk (%) | Risk (%) | Risk difference (%) | Risk ratio |
| Age < 65 years | Death | 1.2 (-0.4-2.8) | 0.3 (-0.1-0.7) | 0.8 (-0.8-2.5) | 3.76 (0.53-26.56) |
|  | Hospitalisation | 16.3 (10.6-21.9) | 11.2 (8.5-13.9) | 5.1 (-1.2-11.3) | 1.45 (0.95-2.22) |
|  | ICU-admission | 1.2 (-0.4-2.8) | 2.5 (1.1-3.8) | -1.3 (-3.4-0.8) | 0.47 (0.11-2.07) |
|  | Mechanical ventilation | 1.2 (-0.4-2.8) | 1.8 (0.7-3.0) | -0.7 (-2.7-1.3) | 0.63 (0.14-2.87) |
|  | Renal replacement therapy | 0.6 (-0.6-1.7) | 1.1 (0.1-2.1) | -0.5 (-2.0-1.0) | 0.54 (0.06-4.67) |
| Age 65+ years | Death | 23.5 (11.8-35.3) | 21.6 (16.0-27.3) | 1.9 (-11.1-14.9) | 1.09 (0.62-1.91) |
|  | Hospitalisation | 60.5 (44.8-76.3) | 54.2 (46.0-62.3) | 6.4 (-11.2-23.9) | 1.12 (0.83-1.51) |
|  | ICU-admission | 18.0 (7.2-28.8) | 10.9 (6.6-15.2) | 7.1 (-4.4-18.6) | 1.65 (0.81-3.37) |
|  | Mechanical ventilation | 15.7 (5.6-25.8) | 9.5 (5.5-13.6) | 6.1 (-4.6-16.9) | 1.64 (0.76-3.53) |
|  | Renal replacement therapy | 3.9 (-1.5-9.3) | 2.9 (0.5-5.2) | 1.1 (-4.8-6.9) | 1.37 (0.28-6.73) |
| Female | Death | 4.5 (1.0-8.0) | 4.8 (2.8-6.8) | -0.3 (-4.4-3.7) | 0.93 (0.38-2.27) |
|  | Hospitalisation | 21.7 (14.6-28.8) | 14.0 (10.5-17.4) | 7.7 (-0.2-15.6) | 1.55 (1.03-2.34) |
|  | ICU-admission | 1.5 (-0.6-3.6) | 1.3 (0.4-2.3) | 0.1 (-2.1-2.4) | 1.11 (0.23-5.29) |
|  | Mechanical ventilation | 1.5 (-0.6-3.6) | 0.6 (-0.1-1.2) | 0.9 (-1.2-3.1) | 2.59 (0.44-15.36) |
|  | Renal replacement therapy | - | 0.8 (0.0-1.5) | - | - |
| Male | Death | 8.9 (3.0-14.8) | 8.0 (5.1-10.9) | 0.9 (-5.7-7.5) | 1.11 (0.52-2.37) |
|  | Hospitalisation | 29.3 (19.0-39.7) | 31.9 (26.3-37.5) | -2.6 (-14.3-9.2) | 0.92 (0.62-1.36) |
|  | ICU-admission | 10.1 (3.8-16.4) | 9.5 (6.2-12.8) | 0.6 (-6.5-7.7) | 1.07 (0.52-2.17) |
|  | Mechanical ventilation | 8.9 (3.0-14.8) | 8.6 (5.4-11.8) | 0.3 (-6.4-7.0) | 1.03 (0.48-2.20) |
|  | Renal replacement therapy | 3.3 (-0.4-7.1) | 2.7 (0.6-4.7) | 0.7 (-3.6-4.9) | 1.25 (0.32-4.83) |
| No history of cardiovascular disease | Death | 4.5 (1.6-7.3) | 3.5 (2.2-4.8) | 1.0 (-2.1-4.1) | 1.29 (0.62-2.69) |
|  | Hospitalisation | 23.9 (17.8-30.1) | 19.3 (16.2-22.5) | 4.6 (-2.3-11.5) | 1.24 (0.91-1.68) |
|  | ICU-admission | 4.5 (1.6-7.3) | 4.1 (2.6-5.7) | 0.4 (-2.9-3.6) | 1.09 (0.52-2.28) |
|  | Mechanical ventilation | 4.0 (1.3-6.7) | 3.5 (2.1-4.9) | 0.5 (-2.6-3.6) | 1.14 (0.51-2.52) |
|  | Renal replacement therapy | 1.0 (-0.4-2.4) | 1.2 (0.3-2.2) | -0.2 (-1.9-1.4) | 0.80 (0.16-3.90) |
| History of cardiovascular disease | Death | 21.7 (4.5-39.0) | 29.7 (19.0-40.4) | -7.9 (-28.0-12.1) | 0.73 (0.31-1.73) |
|  | Hospitalisation | 31.3 (7.8-54.7) | 42.4 (29.2-55.7) | -11.2 (-37.6-15.2) | 0.74 (0.33-1.63) |
|  | ICU-admission | 9.1 (-3.2-21.4) | 10.2 (3.8-16.7) | -1.1 (-14.8-12.5) | 0.89 (0.20-3.86) |
|  | Mechanical ventilation | 8.7 (-3.1-20.5) | 7.9 (2.2-13.5) | 0.8 (-12.1-13.7) | 1.11 (0.24-5.02) |
|  | Renal replacement therapy | 4.3 (-4.2-12.9) | 4.4 (0.1-8.7) | -0.1 (-9.5-9.3) | 0.98 (0.11-8.44) |
| Non-healthcare professional | Death | 7.0 (3.3-10.7) | 8.0 (5.7-10.2) | -0.9 (-5.3-3.4) | 0.88 (0.49-1.60) |
|  | Hospitalisation | 27.9 (21.0-34.7) | 26.4 (22.5-30.4) | 1.4 (-6.5-9.3) | 1.05 (0.79-1.41) |
|  | ICU-admission | 5.4 (2.2-8.7) | 6.1 (4.1-8.1) | -0.7 (-4.5-3.1) | 0.89 (0.45-1.76) |
|  | Mechanical ventilation | 4.9 (1.8-8.0) | 5.1 (3.2-6.9) | -0.2 (-3.8-3.4) | 0.96 (0.46-2.00) |
|  | Renal replacement therapy | 1.1 (-0.4-2.6) | 2.1 (0.8-3.4) | -1.0 (-3.0-1.0) | 0.52 (0.12-2.37) |

\*NSAID use was defined as a filled prescription within 30 days prior to the date of cohort entry.

### Supplementary analyses

Increasing the duration of follow-up to 60 days for death and 30 days for hospitalisation, ICU-admission, mechanical ventilation and acute renal replacement therapy, did not influence the findings (**Supplementary table S2**).

When extending the definition of current NSAID use to a filled prescription up to 60 days before the test date, we observed similar null findings. (**Supplementary table S3**). Likewise, defining exposure as a filled prescription 60 days to 14 days yielded comparable results (**Supplementary table S4**).

### Test negative individuals

We identified 204,920 individuals with a negative SARS-CoV-2 PCR test in the study period. Of these, 3506 were excluded due to migrations within 1 year prior to cohort entry, resulting in a population of 201,414 individuals followed up for a total of 15840 person-years. Use of NSAIDs was associated with a decreased risk of death (RR 0.64, 95% CI 0.49 to 0.84), and increased risk of hospitalisation (RR 1.18, 95% CI 1.08 to 1.28) and ICU-admission (RR 1.39, 95% CI 1.00-1.95) in the adjusted analyses (**Table 4**).

**Table 4:** Association between current NSAID use and 30-day mortality, intensive care unit admission, mechanical ventilation, dialysis and hospitalisation in unmatched and propensity score matched cohorts of individuals tested negative for SARS-CoV-2

| Unmatched cohort |  |  |  |  |  |  |
| --- | --- | --- | --- | --- | --- | --- |
| Outcome | Treated |  | Untreated |  | Comparison |  |
|  | Events | Risk (%) | Events | Risk (%) | Risk difference (%) | Risk ratio |
| Death | 78/5574 | 1.4 (1.1-1.7) | 2830/195840 | 1.4 (1.4-1.5) | -0.0 (-0.4-0.3) | 0.97 (0.77-1.21) |
| Hospitalisation | 757/5091 | 14.9 (13.9-15.8) | 18543/186479 | 9.9 (9.8-10.1) | 4.9 (3.9-5.9) | 1.50 (1.40-1.60) |
| ICU-admission | 63/5547 | 1.1 (0.9-1.4) | 1247/195332 | 0.6 (0.6-0.7) | 0.5 (0.2-0.8) | 1.78 (1.38-2.29) |
| Mechanical ventilation | 38/5558 | 0.7 (0.5-0.9) | 698/195582 | 0.4 (0.3-0.4) | 0.3 (0.1-0.5) | 1.92 (1.38-2.65) |
| Renal replacement therapy | 7/5573 | 0.1 (0.0-0.2) | 166/195754 | 0.1 (0.1-0.1) | 0.0 (-0.1-0.1) | 1.48 (0.70-3.15) |
| Matched cohort |  |  |  |  |  |  |
| Outcome | Treated |  | Untreated |  | Comparison |  |
|  | Events | Risk (%) | Events | Risk (%) | Risk difference (%) | Risk ratio |
| Death | 61/5018 | 1.2 (0.9-1.5) | 382/20072 | 1.9 (1.7-2.1) | -0.7 (-1.1--0.3) | 0.64 (0.49-0.84) |
| Hospitalisation | 651/4613 | 14.1 (13.1-15.1) | 2261/18892 | 12.0 (11.4-12.5) | 2.1 (1.0-3.3) | 1.18 (1.08-1.28) |
| ICU-admission | 50/4992 | 1.0 (0.7-1.3) | 144/20024 | 0.7 (0.6-0.9) | 0.3 (-0.0-0.6) | 1.39 (1.00-1.95) |
| Mechanical ventilation | 29/5003 | 0.6 (0.4-0.8) | 95/20050 | 0.5 (0.4-0.6) | 0.1 (-0.1-0.3) | 1.22 (0.79-1.89) |
| Renal replacement therapy | 6/5017 | 0.1 (0.0-0.2) | 16/20060 | 0.1 (0.0-0.1) | 0.0 (-0.1-0.1) | 1.50 (0.58-3.89) |

\*NSAID use was defined as a filled prescription within 30 days prior to the date cohort entry.
\*\* Patients with a secondary outcome occurring during the outcome assessment window were excluded, resulting in exclusion of n=9844 patients for hospitalisation, n=535 for ICU-admission, n=274 for mechanical ventilation, and n=87 for renal replacement therapy.

## Discussion

We examined whether use of NSAIDs was associated with 30-day mortality and adverse outcomes in a nationwide population of SARS-CoV-2 positive individuals. Use of NSAIDs was not associated with increased 30-day mortality, a finding that was robust in a range of supplementary analyses. Likewise, use of NSAIDs was not associated with an increased risk of hospitalization, ICU-admission, mechanical ventilation, or renal replacement therapy in the adjusted analyses.

The Danish nationwide registries allowed for identification of all individuals who had been tested for SARS-CoV-2 in Denmark and allowed for obtaining data on drug use, medical history, migrations, hospital admissions and death through individual level linkage between health registries. Thus, we were able to include SARS-CoV-2 positive individuals managed in the community as opposed to other data sources that mainly record hospitalised patients. Further, the SARS-CoV-2 test negative population allowed us to conduct a negative control analysis.

In Denmark, NSAIDs can only be obtained via prescription, except for low-dose (200 mg) ibuprofen. With the limited availability and use of NSAIDs over-the-counter in Denmark, misclassification from over-the-counter purchases of NSAIDs is of less concern (18). However, exposure misclassification may still be present since information on adherence, intended duration and dose was not available. Thus, the fact that a patient had filled a prescription for NSAIDs, may be considered an indicator of availability of NSAIDs rather than of actual use of NSAIDs.

The time of cohort entry was defined by the SARS-CoV-2 test date because information on time of symptom onset was not available. Thus, the timing of NSAID use relative to cohort entry will not necessarily reflect NSAID use in the early course of disease.

Users of NSAIDs were more likely to be overweight or obese than non-users both in individuals tested positive and negative for SARS-CoV-2. This is possibly explained by the fact that a diagnosis of overweight or obesity in the Danish National Patient Registry is dependent on a hospital admission or outpatient clinic visit. This is also a likely explanation for the relatively low prevalence of overweight or obesity in this study. The positive predictive value of these diagnoses in the registries is, however, high (27).

Use of NSAIDs has been associated with lower mortality among elderly, presumably because of confounding by indication (28), i.e., NSAIDs are preferentially prescribed to younger, less frail patients because of the established renal-, gastrointestinal-, and cardiovascular adverse effects (29). Considerable media attention of the hypothesized risks with use of NSAIDs in COVID-19, may also have influenced how physicians prescribe NSAIDs to selected patients. Finally, severe symptoms early in the course of disease, before the patient is known to be infected with SARS-CoV-2, may increase the likelihood of being prescribed NSAIDs which would bias the effect estimates towards an increased risk of severe disease associated with NSAIDs.

The secondary outcomes reflecting in-hospital treatment decisions may be more prone to confounding by indication because of the clinical selection of patients to be hospitalised or admitted to the ICU. For example, besides disease severity, factors such as age, comorbidity, and expected outcomes are involved in the ICU triage decision.

Ideally, confounding by indication would be mitigated using an active comparator, however, a suitable active comparator does not exist for ibuprofen. In a previous study, users of paracetamol differed more from NSAID users than non-users of NSAIDs (9).

This study does not provide evidence to withdraw well-indicated use of NSAIDs during the SARS-CoV-2 pandemic. However, the well-established adverse effects of NSAIDs, particularly renal-, gastrointestinal-, and cardiovascular effects should always be considered and NSAIDs should be used in the lowest possible dose for the shortest possible duration for all patients (29).

## Conclusion

Use of NSAIDs was not associated with an increased risk of 30-day mortality or adverse outcomes in patients infected with SARS-CoV-2 in this cohort of all Danish residents tested positive for SARS-CoV-2.

## Data Availability

Due to legal and ethical reasons, individual-level patient data cannot be shared by the authors and is only accessible to authorized researchers after application to the Danish Health Data Authority.

## Acknowledgements

The departments of Clinical Microbiology throughout Denmark are acknowledged for contributing to the national infectious disease surveillance. The Danish Health Data Authority and Statens Serum Institut are acknowledged for valuable support with preparation and linkage of data.

## Contributors and guarantors

All authors designed the study, interpreted the data, revised the manuscript, and approved the final version of the manuscript. KBK and LCL are guarantors.

## Institutional Review Board Approval

The institutional data protection board at the University of Southern Denmark and the Danish Health Data Authority approved the research project. According to Danish law, studies based entirely on registry data do not require approval from an ethics review board (30).

